# Loci on chromosome 12q13.2 encompassing *ERBB3, PA2G4* and *RAB5B* are associated with polycystic ovary syndrome

**DOI:** 10.1101/2022.05.10.22274861

**Authors:** R. Alan Harris, Kellie J. Archer, Mark O. Goodarzi, Timothy P. York, Jeffrey Rogers, Andrea Dunaif, Jan McAllister, Jerome F Strauss

## Abstract

Polycystic ovary syndrome (PCOS) is characterized by hyperandrogenemia of ovarian theca cell origin. Here we report the significant association of 15 single nucleotide polymorphisms (SNPs), identified by whole exome sequencing (WES). DNA was isolated from well-characterized theca cell preparations from women of European ancestry with PCOS (N=9) and elevated androgen production in vitro and from normal ovulatory women (N=7). Of the SNPs, 10 are located within 150 kb on chromosome 12q13.2. This region contains three plausible PCOS candidate genes (*ERBB3/PA2G4/RAB5B*), two of which (*ERBB3* and *RAB5B*) have been identified in GWAS of PCOS. None of the SNPs individually had a significant association with PCOS or in vivo androgen levels when evaluated in an independent cohort (n=318) of families with one or more daughters with PCOS, but a haplotype consisting of the minor alleles of three of the SNPS (rs773121, rs773123 and rs812826) was found preferentially in women with PCOS and elevated androgen levels (p=0.0583). Moreover, the three minor alleles in this haplotype were significantly associated with anti-Mullerian Hormone (AMH) levels, a marker of follicular reserve and follicular maturation. Two of the three SNP minor alleles are predicted to have significant functional consequences (rs773123 a missense SNP in *ERBB3*, and rs812826, a SNP in the *PA2G4* promoter). Notably, *PA2G4* encodes a protein that interacts with the ERBB3 cytoplasmic domain, which is also the domain where the missense variant resides. These findings provide support for the contribution and probable functional significance of loci on chromosome 12q13.2 to the pathophysiology underlying PCOS.

**Author Summary:** Polycystic ovary syndrome (PCOS) is the most common endocrine disorder of women of reproductive age. We identified 15 single nucleotide polymorphisms (SNPs) associated with androgen production in theca cells from normal ovulatory women and women with PCOS. Of these SNPs, 10 are within a 150 kbp region of chromosome 12 including 9 that form a haplotype. This region contains three PCOS candidate genes (*ERBB3/PA2G4/RAB5B*). These SNPs were further examined in an independent cohort of families with one or more daughters with PCOS. A haplotype consisting of the minor alleles of three of the SNPS was found preferentially in women with PCOS. Furthermore, the three minor alleles in this haplotype were significantly associated with anti-Mullerian Hormone (AMH) levels, a marker of follicular reserve and maturation. Two SNPs in the chromosome 12 haplotype were likely to have functional consequences based on genomic context, suggesting that they affect ERBB3 and PA2G4 interactions or *PA2G4* expression.

## Introduction

Polycystic ovary syndrome (PCOS) is the most common endocrine disorder of women of reproductive age. PCOS is characterized by anovulatory infertility, hyperandrogenemia and metabolic disturbance [1]. PCOS is a complex genetic disorder and approximately 20 susceptibility loci have been reproducibly associated in genome wide association studies (GWAS) [2]. However, the functional significance of many of the genes in these loci with respect to PCOS phenotypes is largely unknown [2].

We examined the potential role of genes implicated in PCOS GWAS in controlling androgen production by ovarian theca cells from normal cycling women and women with PCOS, since hyperandrogenemia of ovarian origin is implicated in the pathophysiology of ovarian dysfunction and metabolic disturbances in PCOS [3]. Here we describe SNPs and a haplotype located in a 150 kb region of chromosome 12q13.2 that contains plausible PCOS genes (*ERBB3/PA2G4/RAB5B*), and variants that could have a causal role in promoting PCOS phenotypes.

## Results

### Whole Exome Sequencing Reveals SNPs on Chromosome 12 Associated with Thecal Cell Androgen Production

Table 1 lists the 18 PCOS candidate genes [4–8] interrogated in the whole exome sequencing study. These genes were selected from published GWAS and meta-analyses, representing genes in loci associated with PCOS in women of European ancestry. The variants identified by WES in genes of interest are presented in Supplemental Table S1. The gene locus, chromosome position, rs number, transcript GenBank accession number, the reference and minor alleles, location in the gene, and number of minor alleles detected are provided.

**Table 1:** PCOS Candidate Loci Interrogated.

| **Locus** | **Population** | **GWAS Reference** |
| --- | --- | --- |
| *C9orf3* | Han, European | Shi et al., 2012; Hayes et al., 2015 |
| *DENND1A* | Han, European | Chen et al., 2011; Shi et al., 2012; Day et al., 2018 |
| *ERBB2* | European | Day et al., 2018 |
| *ERBB3* | European | Day et al., 2018 |
| *ERBB4* | European | Day et al., 2015, 2018 |
| *FSHB* | European | Day et al., 2015; Hayes et al., 2015 |
| *GATA4/NEIL2* | European | Hayes et al., 2015 |
| *KRR1* | European | Day et al., 2015, 2018 |
| *MAPRE1* | European | Day et al., 2018 |
| *PLGRKT* | European | Day et al., 2018 |
| *RAB5B/SUOX* | Han, European | Shi et al., 2012; Day et al., 2018 |
| *RAD50* | European | Day et al., 2015, 2018 |
| *THADA* | Han, European | Chen et al., 2011; Shi et al., 2012; Day et al., 2015, 2018 |
| *TOX3* | Han, European | Shi et al., 2012; Day et al., 2018 |
| *YAP1* | Han, European | Shi et al., 2012; Day et al., 2015, 2018 |
| *ZBTB16* | European | Day et al., 2018 |

PCOS theca cells produced significantly more DHEA, the major androgen secreted by theca cell cultures, compared to theca cells from normal ovulatory women under basal conditions, and when challenged with forskolin, which activates adenylate cyclase and mimics the action of luteinizing hormone (LH) (Table 2).

**Table 2:**
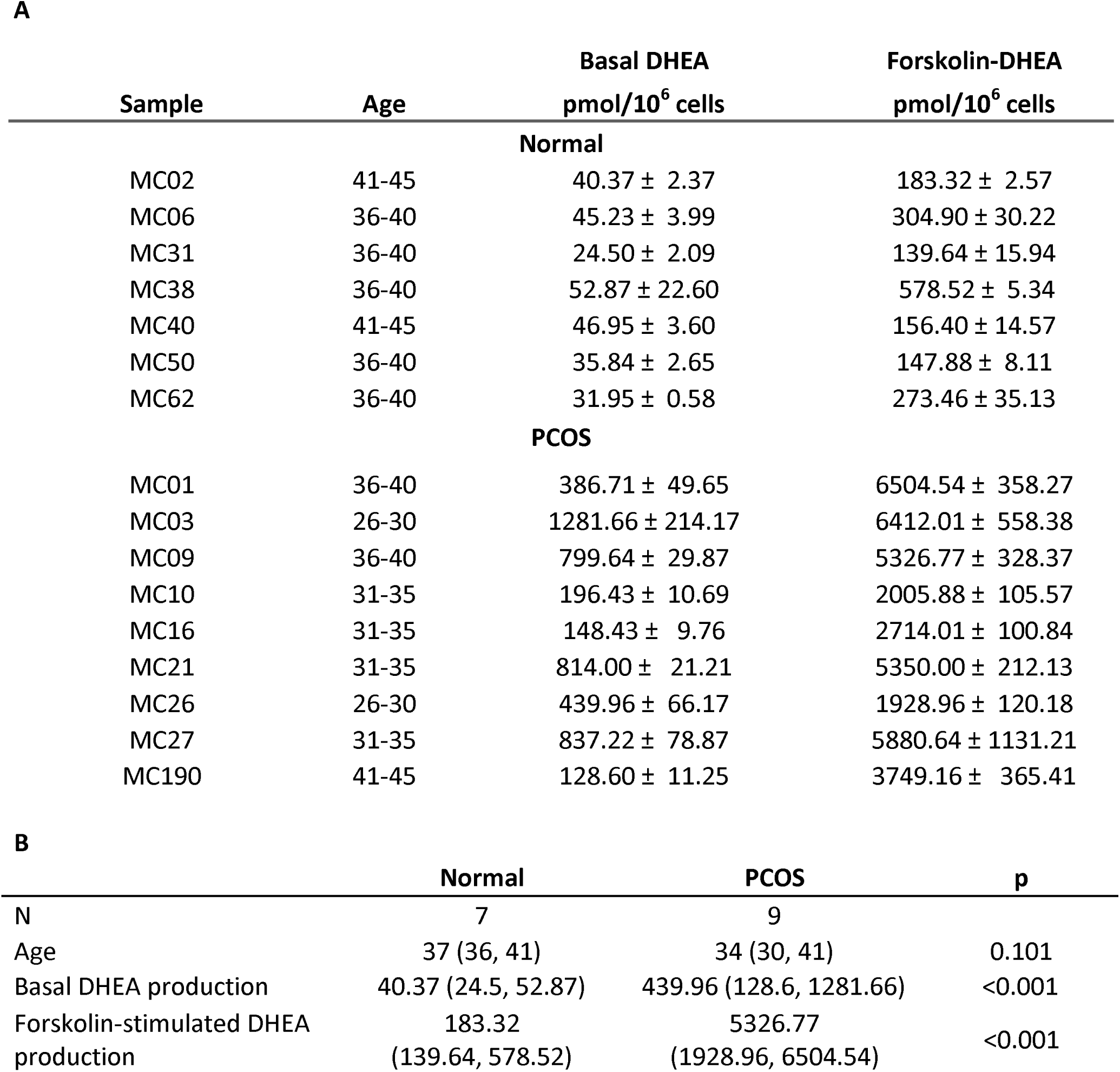
Androgen production by PCOS and normal thecal cells employed in this study. (**A**) DHEA production by normal and PCOS theca cell preparations employed in this study and summary statistics. Cells were cultured for 16 h with or without forskolin (20 μM) and DHEA production was assessed by immunoassay normalized to cell number. Values presented are means (S.D.) from triplicate cultures for each preparation. (**B**) Median and range (minimum, maximum) with P-value from Wilcoxon rank sum test.

Table 3 presents the number of variants analyzed in each gene of interest after filtering to remove duplicate entries and variants that were homogeneous across all cell preparations. An allele-based Wilcoxon rank sum test was then performed on each of the filtered SNPs (N=252) to detect different levels of forskolin-stimulated DHEA production by variant (Table 4).

**Table 3.** Number of variants analyzed by gene after filtering steps applied.

| Gene | Variants |
| --- | --- |
| C9orf3 | 31 |
| DENND1A | 48 |
| ERBB2 | 14 |
| ERBB3 | 23 |
| ERBB4 | 65 |
| FSHB | 4 |
| GATA4 | 42 |
| KRR1 | 17 |
| MAPRE1 | 5 |
| NEIL2 | 15 |
| PLGRKT | 13 |
| RAB5B | 11 |
| RAD50 | 24 |
| SUOX | 4 |
| THADA | 74 |
| TOX3 | 21 |
| YAP1 | 13 |
| ZBTB16 | 17 |

**Table 4.** Variants having significantly different forskolin-stimulated DHEA when comparing samples with the minor allele to those with only the major allele. SNPs significantly associated with thecal androgen production. Nominal p values are presented.

| Gene | Chromosome | Position | dbSNP ID | Major Allele | Minor Allele | P-value |
| --- | --- | --- | --- | --- | --- | --- |
| THADA | chr2 | 43519977 | rs35720761 | C | T | 0.013278 |
| THADA | chr2 | 43736171 | rs6544669 | T | C | 0.007692 |
| ERBB4 | chr2 | 212530466 | rs6725181 | C | T | 0.029670 |
| ERBB4 | chr2 | 212587321 | rs13002712 | G | A | 0.033333 |
| DENND1A | chr9 | 126304695 | rs748994 | T | G | 0.039286 |
| RAB5B | chr12 | 56374318 | rs67594137 | C | T | 0.003571 |
| RAB5B | chr12 | 56374803 | rs11171713 | G | A | 0.003571 |
| RAB5B | chr12 | 56386076 | rs11550558 | A | G | 0.003571 |
| SUOX | chr12 | 56395689 | rs7963590 | G | A | 0.003571 |
| ERBB3 | chr12 | 56473808 | rs7297175 | T | C | 0.041783 |
| ERBB3 | chr12 | 56478607 | rs12817471 | G | A | 0.003571 |
| ERBB3 | chr12 | 56487201 | rs2229046 | T | C | 0.003571 |
| ERBB3 | chr12 | 56494998 | rs773123 | A | T | 0.003571 |
| ERBB3 | chr12 | 56495306 | rs812826 | C | T | 0.003571 |
| ERBB3 | chr12 | 56498241 | rs773121 | G | A | 0.003571 |

Among the 15 variants that were found to be significant at a *P*-value < 0.05, 10 were located on chromosome 12. This over representation of chromosome 12 was highly significant (Fisher’s exact test, P=0.000002).

The variants on chromosome 12 showing significant effects include 10 SNPs, 6 in the *ERBB3* gene (rs7297125, rs12817471, rs2229046, rs773123, rs812826, rs773121), and 4 (rs67594137, rs11171713, rs11550558, rs7963590) in *RAB5B/SUOX*. The latter genes overlap each other on opposite DNA strands. Notably, the same PCOS theca cell preparations (MC01, MC03, MC27 representing one third of the PCOS sample) contained the minor allele of these SNPs in a heterozygous state, suggesting linkage disequilibrium (LD) and a large effect size. The LD was not unexpected since the variants are located within a 150 kb stretch of chromosome 12q13.2. Moreover, using LDlink programs to investigate linkage disequilibrium in European populations, we found that one SNP, rs7297175, is independent of the other 9, while 6 SNPs (rs67594137, rs11171713, rs11550558, rs7963590, rs12817471, rs2229046) formed one LD group (r^2^ 0.77 to 0.97 among the SNPs) and 3 SNPs (rs773123, rs812826, rs773121) formed another LD group (r^2^ 1.0) (Figure 1). We then used LDlink to identify the haplotypes present in the two haplotype blocks, finding two haplotypes in each block, one consisting of the major allele of the constituent SNPs and the other containing the minor alleles (Figure 2).

**Fig 1.**
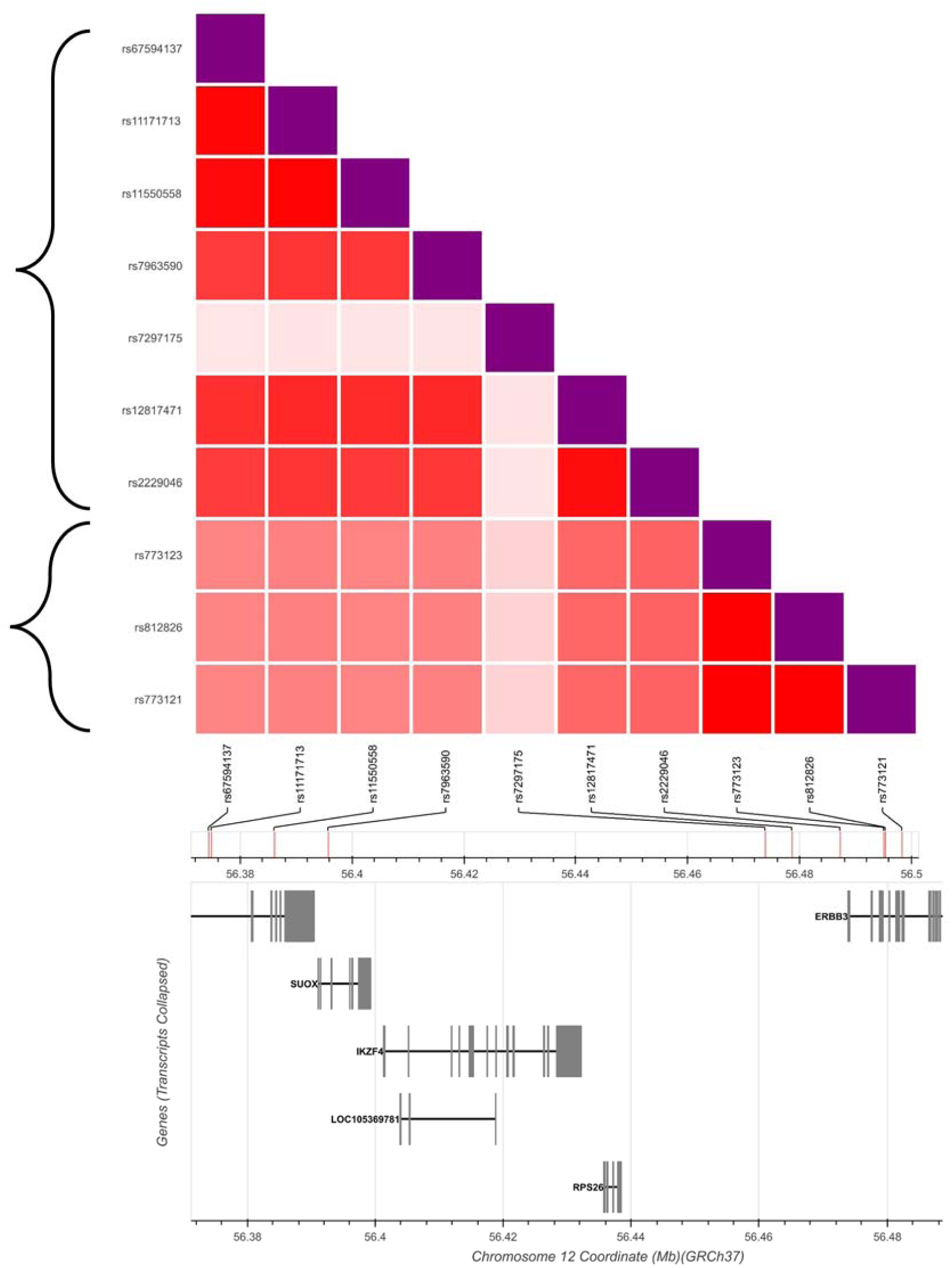
Linkage disequilibrium among chromosome 12 SNPs in Individuals of European Ancestry. The plot displays linkage disequilibrium as r2 between each pair of SNPs. The brackets outline the two linkage disequilibrium groups (r2 > 0.5). Note that rs7297175 is independent of the other 9 SNPs and is not included in either haplotype block. Darker red indicates higher linkage disequilibrium. SNP locations and genes in the region are displayed at bottom.

**Fig 2.**
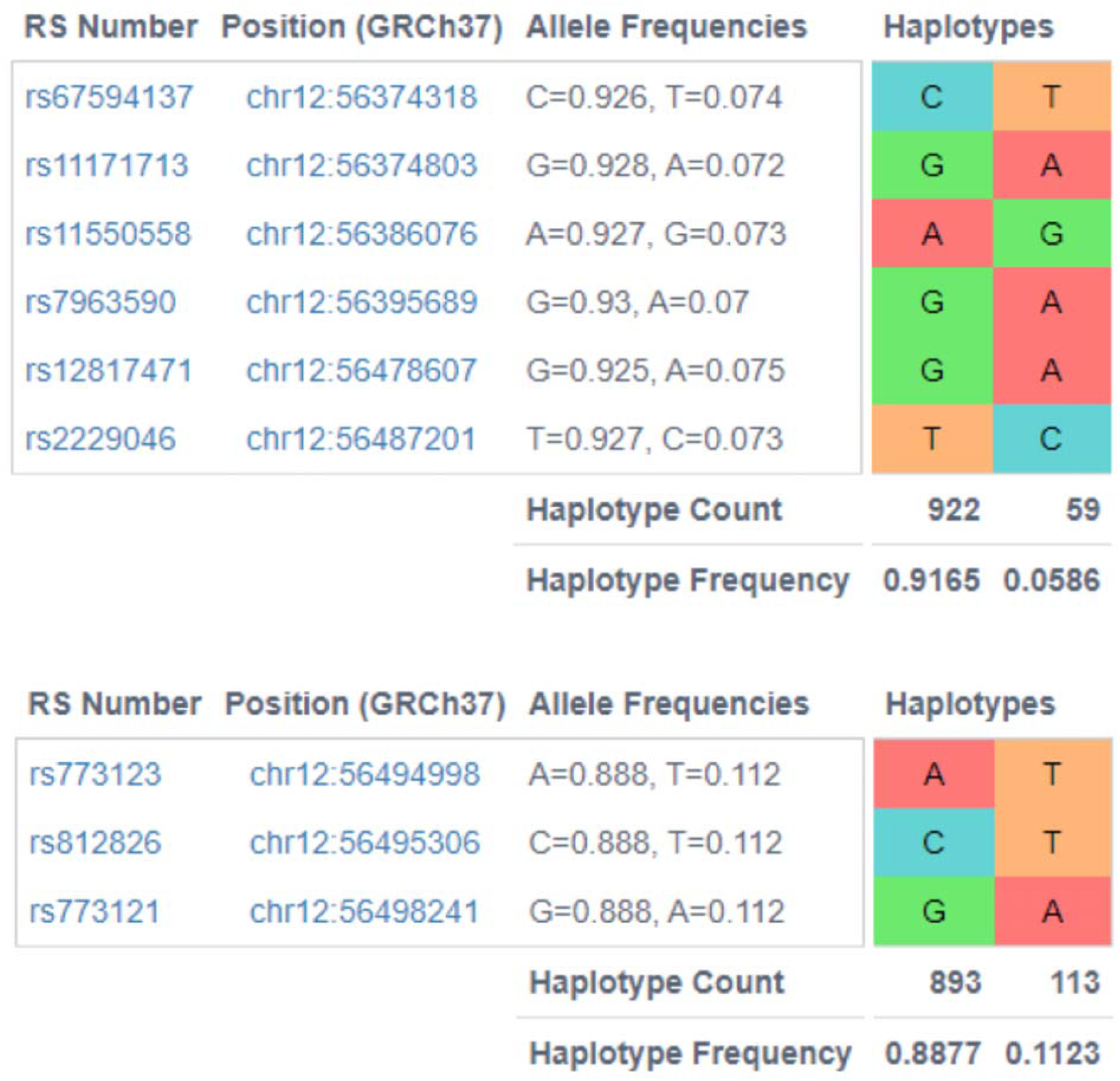
Haplotypes in the chromosome 12 region of interest. The first haplotype block consists of 6 SNPs and the second consists of 3 SNPs. Each block contains a common haplotype and a rare haplotype.

### Analysis of Whole Genome Sequences from a Cohort of European Ancestry Women

We examined the chromosome 12 SNPs in whole genome sequencing data from a cohort of 318 individuals of European ancestry from 77 families with one or more daughters with PCOS [9]. PLINK [10] transmission disequilibrium tests (TDT) on the individual SNPs did not identify any of them as significantly associated with PCOS/HA (hyperandrogenemia) affection status [11] (Table S2). However, rs773123, rs812826, and rs773121, corresponding to one of the haplotype groups identified in the initial samples, had p values ranging from 0.1011 to 0.1404 compared to p values ranging from 0.4652 to 0.8575 for the other SNPs. Based on the lower range of p values for rs773123, rs812826, and rs773121, we examined the possibility they formed a haplotype with association to PCOS/HA using the Family-Based Association Tests (FBAT) [12] HBAT test which is the haplotype version of the association test. Based on the HBAT test, the haplotype consisting of the minor allele for rs773123, rs812826, and rs773121 and the major allele for the remaining SNPs was found preferentially in women with PCOS and elevated androgen levels (p = 0.0583) (Table S3).

Association analyses of levels of total testosterone (T), DHEAS (dehydroepiandrosterone sulfate), sex hormone binding globulin (SHBG), luteinizing hormone (LH), follicle stimulating hormone (FSH), and anti-Mullerian hormone (AMH) were performed using the PLINK family-based association test for quantitative traits (QFAM). After 100,000 permutations for empirical p-value correction of QFAM total results, the only associations significant at p < 0.01 were AMH associations with rs773123 (P_emp_ = 0.00105), rs812826 (P_emp_ = 0.0009), and rs773121 (P_emp_ = 0.00242). The minor alleles of these same SNPs were present in the haplotype that approached significance in the affection status FBAT HBAT test. The quantitative trait data for T, DHEAS, SHBG, LH and FSH were reported as part of our previous study using family-based WGS analyses [9]. The AMH median (25th to 75th percentiles) value (ng/mL) for affected women was 10.02 (6.64-14.31) and for unaffected women, it was 3.02 (1.06-4.77). No quantitative analyses of hormone levels using FBAT HBAT were significant.

### Functional Annotations of SNPs

To examine the potential functional impact of the chromosome 12 SNPs, we annotated them with CADD PHRED scores [13]. A CADD PHRED score of 10 predicts a SNP is among the 10% most functional changes in the human genome while a CADD PHRED score of 20 indicates the change is among the 1% most functional changes. Three of the SNPs have a CADD PHRED score greater than 10 including two in the HBAT identified haplotype (Table S4). rs773123 (CADD PHRED 24.6) is a missense SNP in *ERBB3* at amino acid position 1119 which is in the cytoplasmic topological domain (https://www.uniprot.org/uniprot/P21860#subcellular_location). rs773121 (CADD PHRED 13.2) is in the promoter (Ensembl regulatory feature ENSR00000052477) for *PA2G4* which is a transcriptional co-repressor of androgen receptor-regulated genes [14]. PA2G4 interacts with the cytoplasmic domain of ERBB3 [15] which contains the rs773123 missense SNP. The *PA2G4* Ensembl regulatory feature ENSR00000052477 is equivalent to the GeneHancer [16] GH12J056102 regulatory element. This GeneHancer regulatory element is classified as having both promoter and enhancer functions and two of the genes for which it has a predicted enhancer function are *ERBB3* and *RAB5B*. These interactions can be viewed in the UCSC Genome Browser “Interactions between GeneHancer regulatory elements and genes” track (https://genome.ucsc.edu/s/Rharris1/chr12.PCOS). Since these two SNPs occur in a haplotype with a p value approaching significance, there could be some interplay between SNP rs773121’s regulatory effect on the expression of *PA2G4* and *ERBB3* and SNP rs773123’s effect on the interaction between the ERBB3 cytoplasmic domain and PA2G4. Additionally, *PA2G4* and *ERBB3* are targets of transcription factor, *ZNF217*, another PCOS candidate gene identified by GWAS (https://maayanlab.cloud/Harmonizome/gene_set/ZNF217/ENCODE+Transcription+Factor+Targets) [17].

## Discussion

We found evidence of an association between androgen (DHEA) production by cultures of human theca cells, a reflection of theca cell endocrine activity, and fifteen SNPs, including ten in a haplotype on chromosome 12. This haplotype was found preferentially in women with PCOS and elevated androgen levels (p = 0.0583) in an independent family cohort. Two minor alleles in the chromosome 12 haplotype were likely to have functional consequences based on CADD scores and genomic context, suggesting that they affect ERBB3 and PA2G4 interactions (rs773123) or *PA2G4* expression (rs773121). Moreover, a recent study [18] utilizing a different methodologic approach, colocalization analysis, identified the same region on chromosome 12 encompassing *ERBB3, PA2G4, RAB5B* and *SUOX*, as containing potential PCOS disease-mediating genes.

ERBB3 is a component of the EGF family of signal transduction factors. It forms heterodimers with other ERBB family members, including ERBB2, which is the receptor for neuregulins, which are produced by both theca cells and granulosa cells in response to LH [19]. Neuregulin-1 (NRG-1) also plays a role in the proliferation of Leydig cells, an androgen producing cell type in the male gonad that is functionally analogous to theca cells in the female gonad. Thus, the haplotype identified in this report encompasses genes that are implicated in the regulation/function of androgen producing cells. Moreover, both *ERBB3* and *RAB5B* have been associated with metabolic phenotypes that are related to PCOS, including glucose metabolism/insulin resistance [4,5,20].

GWAS conducted on Han Chinese identified *RAB5B/SUOX* as PCOS candidate loci [7]. The minor alleles of the SNPs we identified have very different allele frequencies in non-Finnish European and East Asian populations with the exception of rs11550558, being low frequency in East Asians (<0.05%) and 7-11% in Europeans (Table S5). A recent case-control study of SNPs encoding the 3’-UTR of the *RAB5B* gene conducted in Han Chinese revealed highly significant associations with PCOS phenotypes with large effect sizes [21]. These findings support our conclusions that 12q13.2 contains important genetic determinants of PCOS, and that the specific SNPs that influence thecal androgen production are population-specific. The SNPs may impact the level of expression of the candidate genes, accounting for variation in cell/tissue function, including the endocrine functions of thecal cells and granulosa cells. For example, ERBB3 is expressed by granulosa cells [19], which produce AMH, a peptide factor that influences growth of follicles [22]. Circulating AMH levels are elevated in women with PCOS, suggesting that the chromosome 12 haplotype plays a role in regulating both theca cell (androgen) and granulosa (AMH) cell function [23].

RAB5B is involved in intracellular trafficking of endosomes including those derived from the plasma membrane [24]. We have previously shown that RAB5B is co-localized in compartments containing DENND1A.V2, another PCOS GWAS candidate gene associated with hyperandrogenemia [25,26]. DENND1A.V2 has an N-terminal guanine nucleotide exchange function and a clathrin-binding domain, putting it at the nexus with plasma membrane signaling proteins like the ERBB family of proteins. DENND1A.V2 is translocated into the nucleus of PCOS theca cells along with RAB5B, suggesting a role in regulation of expression of genes involved in androgen synthesis [26]. *SUOX*, which overlaps the *RAB5B* gene, encodes a mitochondrial sulfite oxidase, which has not been linked to PCOS, thecal cell function or steroidogenesis. Thus, SNPs in this gene are likely to be irrelevant to the PCOS phenotypes.

In summary, our findings provide support for functional contributions of genes on chromosome 12q13.2, including *ERBB3, PA2G4*, and *RAB5B*, to ovarian cell dysfunction in PCOS. Our findings align with in silico colocalization analyses implicating these same genes in PCOS pathogenesis.

## Materials and Methods

### Theca cell preparations and culture

Human theca interna tissue was obtained from follicles of women undergoing hysterectomy, following informed consent under a protocol approved by the Institutional Review Board of The Pennsylvania State University College of Medicine. As a standard of care, oophorectomies were performed during the luteal phase of the cycle. Theca cells from normal cycling and PCOS follicles were isolated and grown as we have as previously reported in detail[27–29]. PCOS and normal ovarian tissue came from age-matched women, 38–41 years old. The diagnosis of PCOS was made according to National Institutes of Health (NIH) consensus guidelines [30,31] which include hyperandrogenemia/hyperandrogenism and oligo-ovulation and the exclusion of other causes of hyperandrogenemia (e.g. 21-hydroxylase deficiency, Cushing’s syndrome, and adrenal or ovarian tumors). All of the PCOS theca cell preparations studied came from ovaries of women with fewer than six menses per year and elevated serum total testosterone or bioavailable testosterone levels [32–35]. Each of the PCOS ovaries contained multiple subcortical follicles of less than 10 mm in diameter. The control (normal) theca cell preparations came from ovaries of fertile women with normal menstrual histories, menstrual cycles of 21–35 days, and no clinical signs of hyperandrogenism. Neither PCOS nor normal subjects were receiving hormonal medications at the time of surgery. Indications for surgery were dysfunctional uterine bleeding, endometrial cancer, and pelvic pain. Experiments comparing PCOS and normal theca were performed using fourth-passage (31–38 population doublings) theca cells isolated from individual size-matched follicles obtained from age-matched subjects, in the absence of in vivo stimulation. The use of fourth-passage cells allowed us to perform multiple experiments from the same patient population, and were propagated from frozen stocks of second passage cells in the media described above. The passage conditions and split ratios for all normal and PCOS cells were identical. These studies were approved by the Human Subjects Protection Offices of Virginia Commonwealth University) and Penn State College of Medicine.

Nine cell preparations obtained from women with PCOS (MC01, MC21, MC09_B, MC03, MC10, MC16, MC26, MC27, MC190) and seven from normal ovulating women (MC62_B, MC02, MC06, MC31, MC38, MC40, MC50) were studied. All subjects were unrelated and of European ancestry. The cells were characterized by their production of dehydroepiandrosterone (DHEA), the major androgen synthesized by these cells, under basal conditions or stimulated with forskolin (20 μM) for 16 h. DHEA was quantified by ELISA assays (DRG, Springfield,NJ) and production (pmol) was normalized to cell number (10^6^ cells) determined at the end of the culture period.

### Whole Exome Sequence Analysis of Normal and PCOS Theca Cells DNA

The DNA samples were subjected to whole exome sequencing at 100 millions reads providing 100× coverage using the Agilent SureSelect 51M capture kit with Illumina HiSeq 2000 sequencing, in conjunction with BGI Americas. Raw sequence data for each individual were mapped to the human reference genome (build GRCh37/hg19) using the BWA-MEM algorithm of Burrows-Wheeler Aligner (v 0.7.12) (H. Li, 2013). This was followed by a series of pre-processing steps–marking duplicates, realignment around indels and base quality recalibration. PCR duplicates were marked within the aligned reads using Picard tools. (http://picard.sourceforge.net) Next, mapping artifacts around indels were cleaned up using the RealignerTargetCreator, the IndelRealigner and the LeftAlignIndels walkers of the Genome Analysis ToolKit (GATK) [37,38]. Inaccurate / biased base quality scores were recalibrated using the BaseRecalibrator, the AnalyzeCovariates and the PrintReads walkers of GATK, which use machine learning to model these errors empirically and adjust the quality scores accordingly.

### Linkage disequilibrium

LDlink (https://analysistools.cancer.gov/LDlink/?tab=home) was used to identify haplotypes in Europeans. LDlink accesses data from 1000 Genomes in a suite of tools that allows determination of linkage disequilibrium (LD) and haplotypes. We used the LDlink SNPclip tool to examine LD among the 9 SNPs associated with DHEA response, using an r2 cutoff of 0.5. This identified two LD groups, which were then used to identify haplotypes using LDhap.

### Statistical analysis

The Wilcoxon rank sum test was used to compare age, basal DHEA, and forskolin-stimulated DHEA production between the PCOS and control samples. The WES data were subjected to the following filters. We retained genetic variants having a unique combination of Gene ID, Chromosome, Position, Variant ID, Reference and Alternate Allele. Additionally, variants that were homogeneous across all samples (i.e., no sample displayed the minor allele or all samples displayed the minor allele (N=21)) were removed, leaving 441 variants for statistical analysis.

For each variant, a Wilcoxon rank sum test was used to compare those with and without the variant with respect to forskolin-stimulated DHEA production. In order to apply a statistical comparison, a minimum of two samples per group were required so that the set of variants was restricted to 252 variants. P values of <0.05 were considered significant. We did not apply Bonferroni correction because we are testing a restricted set of genetic variants in robust loci for PCOS.

### SNP analyses of a PCOS cohort

We analyzed the 10 SNPs on chromosome 12 that were significantly associated with thecal cell androgen production in whole genome sequencing data from a family based PCOS cohort [9]. The study was approved by the Institutional Review Boards of Northwestern University Feinberg School of Medicine, Penn State Health Milton S. Hershey Medical Center, and Brigham and Women’s Hospital. Written informed consent was obtained from all subjects prior to the study. The cohort consisted of 318 individuals of European ancestry from 77 families with one or more daughters with PCOS. Among the index cases and sisters (n=171), the following phenotypes were identified: PCOS (T>58 ng/dl and/or uT>15 ng/dl and ≤ 8 menses/year) (n=90); Hyperandrogenemic (HA) (T>58 ng/dl and/or uT>15 ng/dl and regular menses (every 27-35 days)) (n=5); Unaffected (n=76). The women were ages 14 to 49 years. Women were assigned affected status if they fulfilled criteria for PCOS or HA, as we have done in our previous family-based genetic analyses [11]. All anthropometric and hormonal data, except for circulating AMH levels, have been previously reported [9].

Sequencing of the cohort was performed using the Complete Genomics, Inc. platform. Sequence reads were aligned to the human reference genome (GRCh37/hg19) and variants were called using the CGI AssemblyPipeline version 2.0. The SNPs were analyzed individually using the PLINK v1.90 [10] transmission disequilibrium test (TDT) based on PCOS affection status. An individual was considered affected if they had a phenotype of PCOS or Hyperandrogenic. The PLINK family-based association test for quantitative traits (QFAM) was used to examine associations of quantitative traits with the SNPs measured in the index cases and sisters. The sample size was T (n=162), DHEAS (n=161), SHBG (n=160), LH (n=162) and FSH (n=162). There was insufficient sample for AMH assays in some subjects, thus, the sample size with AMH values was smaller (n=59). QFAM total results were empirically corrected using 100,000 permutations. The haplotypes containing the SNPs were analyzed using the Family-Based Association Tests (FBAT) v2.0.3 [12] HBAT function which is the haplotype version of the association test. FBAT HBAT was performed using both the PCOS affection status and quantitative traits. Potential functional consequences of the SNPs were examined using Combined Annotation-Dependent Depletion (CADD) v1.6 [13] and GeneHancer [16].

## Data Availability

All data produced in the present work are contained in the manuscript

## Supporting information captions

### Supplemental Table 1.xlsx

**Table S1. WES results for selected PCOS candidate genes**. The table presents the gene name, chromosome assignment, nucleotide position of the detected variant, rs number, major and minor alleles, location and/or variant effect on coding sequence, transcript, and distribution of minor alleles among the different theca cell preparations designated by their MC number (see below), with number of homozygous (left), heterozygous (middle) followed by the total number of minor alleles (right) detected.

### Supplemental.docx

**Table S2**. PLINK transmission disequilibrium tests (TDT) based on affection status for the individual chromosome 12 SNPs.

**Table S3**. Family-Based Association Tests (FBAT) HBAT test based on affection status for chromosome 12 SNPs.

**Table S4**. CADD PHRED scores and a subset of annotation details for chromosome 12 SNPs.

**Table S5. Minor Allele Frequencies for the 12q13.2 SNPs**. Minor allele frequencies for non-Finnish Europeans, East Asians and African Americans were extracted from GnomAD (https://gnomad.broadinstitute.org). rs7297175 is independent of the other 9 SNPs and is not included in either haplotype block.

## References

1. Diamanti-Kandarakis E, Dunaif A. Insulin resistance and the polycystic ovary syndrome revisited: an update on mechanisms and implications. Endocr Rev. 2012;33: 981–1030. doi:10.1210/ER.2011-1034

2. Dapas M, Dunaif A. Deconstructing a Syndrome: Genomic Insights into PCOS Causal Mechanisms and Classification. Endocr Rev. 2022 [cited 19 Apr 2022]. doi:10.1210/ENDREV/BNAC001

3. Legro RS, Driscoll D, Strauss JF, Fox J, Dunaif A. Evidence for a genetic basis for hyperandrogenemia in polycystic ovary syndrome. Proc Natl Acad Sci U S A. 1998;95: 14956–14960. doi:10.1073/PNAS.95.25.14956

4. Day FR, Hinds DA, Tung JY, Stolk L, Styrkarsdottir U, Saxena R, et al. Causal mechanisms and balancing selection inferred from genetic associations with polycystic ovary syndrome. Nat Commun. 2015;6. doi:10.1038/NCOMMS9464

5. Day F, Karaderi T, Jones MR, Meun C, He C, Drong A, et al. Large-scale genome-wide meta-analysis of polycystic ovary syndrome suggests shared genetic architecture for different diagnosis criteria. PLoS Genet. 2018;14. doi:10.1371/JOURNAL.PGEN.1007813

6. Hayes MG, Urbanek M, Ehrmann DA, Armstrong LL, Lee JY, Sisk R, et al. Genome-wide association of polycystic ovary syndrome implicates alterations in gonadotropin secretion in European ancestry populations. Nat Commun. 2015/08/19. 2015;6: 7502. doi:10.1038/ncomms8502

7. Shi Y, Zhao H, Shi Y, Cao Y, Yang D, Li Z, et al. Genome-wide association study identifies eight new risk loci for polycystic ovary syndrome. Nat Genet. 2012;44: 1020–1025. doi:10.1038/NG.2384

8. Chen ZJ, Zhao H, He L, Shi Y, Qin Y, Shi Y, et al. Genome-wide association study identifies susceptibility loci for polycystic ovary syndrome on chromosome 2p16.3, 2p21 and 9q33.3. Nat Genet. 2011;43: 55–59. doi:10.1038/NG.732

9. Dapas M, Sisk R, Legro RS, Urbanek M, Dunaif A, Hayes MG. Family-based quantitative trait meta-analysis implicates rare noncoding variants in DENND1A in polycystic ovary syndrome. J Clin Endocrinol Metab. 2019;104: 3835–3850. doi:10.1210/JC.2018-02496

10. Purcell S, Neale B, Todd-Brown K, Thomas L, Ferreira MAR, Bender D, et al. PLINK: a tool set for whole-genome association and population-based linkage analyses. Am J Hum Genet. 2007;81: 559–575. doi:10.1086/519795

11. Urbanek M, Legro RS, Driscoll DA, Azziz R, Ehrmann DA, Norman RJ, et al. Thirty-seven candidate genes for polycystic ovary syndrome: strongest evidence for linkage is with follistatin. Proc Natl Acad Sci U S A. 1999;96: 8573–8578. doi:10.1073/PNAS.96.15.8573

12. Horvath S, Xu X, Laird NM. The family based association test method: strategies for studying general genotype--phenotype associations. Eur J Hum Genet. 2001;9: 301–306. doi:10.1038/SJ.EJHG.5200625

13. Rentzsch P, Witten D, Cooper GM, Shendure J, Kircher M. CADD: predicting the deleteriousness of variants throughout the human genome. Nucleic Acids Res. 2019;47: D886–D894. doi:10.1093/NAR/GKY1016

14. Zhang Y, Akinmade D, Hamburger AW. The ErbB3 binding protein Ebp1 interacts with Sin3A to repress E2F1 and AR-mediated transcription. Nucleic Acids Res. 2005;33: 6024–6033. doi:10.1093/NAR/GKI903

15. Yoo JY, Wang XW, Rishi AK, Lessor T, Xia XM, Gustafson TA, et al. Interaction of the PA2G4 (EBP1) protein with ErbB-3 and regulation of this binding by heregulin. Br J Cancer. 2000;82: 683–690. doi:10.1054/BJOC.1999.0981

16. Fishilevich S, Nudel R, Rappaport N, Hadar R, Plaschkes I, Stein TI, et al. GeneHancer: genome-wide integration of enhancers and target genes in GeneCards. Database (Oxford). 2017;2017. doi:10.1093/DATABASE/BAX028

17. Krig SR, Miller JK, Frietze S, Beckett LA, Neve RM, Farnham PJ, et al. ZNF217, a candidate breast cancer oncogene amplified at 20q13, regulates expression of the ErbB3 receptor tyrosine kinase in breast cancer cells. Oncogene. 2010;29: 5500–5510. doi:10.1038/ONC.2010.289

18. Censin JC, Bovijn J, Holmes M v, Lindgren CM. Colocalization analysis of polycystic ovary syndrome to identify potential disease-mediating genes and proteins. Eur J Hum Genet. 2021/03/06. 2021;29: 1446–1454. doi:10.1038/s41431-021-00835-8

19. Chowdhury I, Branch A, Mehrabi S, Ford BD, Thompson WE. Gonadotropin-Dependent Neuregulin-1 Signaling Regulates Female Rat Ovarian Granulosa Cell Survival. Endocrinology. 2017;158: 3647–3660. doi:10.1210/EN.2017-00065

20. Jones MR, Brower MA, Xu N, Cui J, Mengesha E, Chen YDI, et al. Systems Genetics Reveals the Functional Context of PCOS Loci and Identifies Genetic and Molecular Mechanisms of Disease Heterogeneity. PLoS Genet. 2015;11. doi:10.1371/JOURNAL.PGEN.1005455

21. Yu J, Ding C, Guan S, Wang C. Association of single nucleotide polymorphisms in the RAB5B gene 3’UTR region with polycystic ovary syndrome in Chinese Han women. Biosci Rep. 2019/05/01. 2019;39. doi:10.1042/BSR20190292

22. Dumont A, Robin G, Catteau-Jonard S, Dewailly D. Role of Anti-Müllerian Hormone in pathophysiology, diagnosis and treatment of Polycystic Ovary Syndrome: a review. Reprod Biol Endocrinol. 2015;13. doi:10.1186/S12958-015-0134-9

23. Gorsic LK, Kosova G, Werstein B, Sisk R, Legro RS, Hayes MG, et al. Pathogenic Anti-Müllerian Hormone Variants in Polycystic Ovary Syndrome. J Clin Endocrinol Metab. 2017;102: 2862–2872. doi:10.1210/JC.2017-00612

24. Gulappa T, Clouser CL, Menon KM. The role of Rab5a GTPase in endocytosis and post-endocytic trafficking of the hCG-human luteinizing hormone receptor complex. Cell Mol Life Sci. 2011;68: 2785–2795. doi:10.1007/s00018-010-0594-1

25. McAllister JM, Legro RS, Modi BP, Strauss 3rd JF. Functional genomics of PCOS: from GWAS to molecular mechanisms. Trends Endocrinol Metab. 2015/01/21. 2015;26: 118–124. doi:10.1016/j.tem.2014.12.004

26. Kulkarni R, Teves ME, Han AX, McAllister JM, Strauss 3rd JF. Colocalization of Polycystic Ovary Syndrome Candidate Gene Products in Theca Cells Suggests Novel Signaling Pathways. J Endocr Soc. 2019/11/15. 2019;3: 2204–2223. doi:10.1210/js.2019-00169

27. Wickenheisser JK, Nelson-Degrave VL, McAllister JM. Dysregulation of cytochrome P450 17alpha-hydroxylase messenger ribonucleic acid stability in theca cells isolated from women with polycystic ovary syndrome. J Clin Endocrinol Metab. 2005;90: 1720–1727. Available: http://www.ncbi.nlm.nih.gov/entrez/query.fcgi?cmd=Retrieve&db=PubMed&dopt=Citation&list_uids=15598676

28. Wickenheisser JK, Biegler JM, Nelson-Degrave VL, Legro RS, Strauss 3rd JF, McAllister JM. Cholesterol side-chain cleavage gene expression in theca cells: augmented transcriptional regulation and mRNA stability in polycystic ovary syndrome. PLoS One. 2012/11/17. 2012;7: e48963. doi:10.1371/journal.pone.0048963

29. Nelson-Degrave VL, Wickenheisser JK, Hendricks KL, Asano T, Fujishiro M, Legro RS, et al. Alterations in mitogen-activated protein kinase kinase and extracellular regulated kinase signaling in theca cells contribute to excessive androgen production in polycystic ovary syndrome. Mol Endocrinol. 2005;19: 379–390. Available: http://www.ncbi.nlm.nih.gov/entrez/query.fcgi?cmd=Retrieve&db=PubMed&dopt=Citation&list_uids=15514033

30. Legro RS, Arslanian SA, Ehrmann DA, Hoeger KM, Murad MH, Pasquali R, et al. Diagnosis and treatment of polycystic ovary syndrome: an endocrine society clinical practice guideline. J Clin Endocrinol Metab. 2013/10/24. 2013;98: 4565–4592. doi:10.1210/jc.2013-2350

31. Azziz R, Carmina E, Chen Z, Dunaif A, Laven JS, Legro RS, et al. Polycystic ovary syndrome. Nat Rev Dis Primers. 2016/08/12. 2016;2: 16057. doi:10.1038/nrdp.2016.57

32. Nelson-DeGrave VL, Wickenheisser JK, Cockrell JE, Wood JR, Legro RS, Strauss 3rd JF, et al. Valproate potentiates androgen biosynthesis in human ovarian theca cells. Endocrinology. 2004;145: 799–808. Available: http://www.ncbi.nlm.nih.gov/entrez/query.fcgi?cmd=Retrieve&db=PubMed&dopt=Citation&list_uids=14576182

33. Nelson VL, Legro RS, Strauss 3rd JF, McAllister JM. Augmented androgen production is a stable steroidogenic phenotype of propagated theca cells from polycystic ovaries. Mol Endocrinol. 1999;13: 946–957. Available: http://www.ncbi.nlm.nih.gov/entrez/query.fcgi?cmd=Retrieve&db=PubMed&dopt=Citation&list_uids=10379893

34. Wickenheisser JK, Nelson-DeGrave VL, Quinn PG, McAllister JM. Increased cytochrome P450 17alpha-hydroxylase promoter function in theca cells isolated from patients with polycystic ovary syndrome involves nuclear factor-1. Mol Endocrinol. 2004;18: 588–605. Available: http://www.ncbi.nlm.nih.gov/entrez/query.fcgi?cmd=Retrieve&db=PubMed&dopt=Citation&list_uids=14684846

35. Wickenheisser JK, Quinn PG, Nelson VL, Legro RS, Strauss 3rd JF, McAllister JM. Differential activity of the cytochrome P450 17alpha-hydroxylase and steroidogenic acute regulatory protein gene promoters in normal and polycystic ovary syndrome theca cells. J Clin Endocrinol Metab. 2000;85: 2304–2311. Available: http://www.ncbi.nlm.nih.gov/entrez/query.fcgi?cmd=Retrieve&db=PubMed&dopt=Citation&list_uids=10852468

36. Li H. Aligning sequence reads, clone sequences and assembly contigs with BWA-MEM. 2013 [cited 19 Dec 2017]. Available: http://arxiv.org/abs/1303.3997

37. McKenna A, Hanna M, Banks E, Sivachenko A, Cibulskis K, Kernytsky A, et al. The Genome Analysis Toolkit: a MapReduce framework for analyzing next-generation DNA sequencing data. Genome Res. 2010;20: 1297–1303. doi:10.1101/GR.107524.110

38. Depristo MA, Banks E, Poplin R, Garimella K v., Maguire JR, Hartl C, et al. A framework for variation discovery and genotyping using next-generation DNA sequencing data. Nat Genet. 2011;43: 491–501. doi:10.1038/NG.806

