## Supplemental Tables 2-5 for "Loci on chromosome 12q13.2 encompassing *ERBB3, PA2G4* and *RAB5B* are associated with polycystic ovary syndrome"

| **CHR** | **SNP** | **BP** | **A1** | **A2** | **T** | **U** | **OR** | **CHISQ** | **P** |
| --- | --- | --- | --- | --- | --- | --- | --- | --- | --- |
| 12 | rs67594137 | 56374318 | T | C | 14 | 17 | 0.8235 | 0.2903 | 0.59 |
| 12 | rs11171713 | 56374803 | A | G | 13 | 17 | 0.7647 | 0.5333 | 0.4652 |
| 12 | rs11550558 | 56386076 | G | A | 14 | 17 | 0.8235 | 0.2903 | 0.59 |
| 12 | rs7963590 | 56395689 | A | G | 15 | 16 | 0.9375 | 0.03226 | 0.8575 |
| 12 | rs12817471 | 56478607 | A | G | 15 | 17 | 0.8824 | 0.125 | 0.7237 |
| 12 | rs2229046 | 56487201 | C | T | 15 | 17 | 0.8824 | 0.125 | 0.7237 |
| 12 | rs773123 | 56494998 | T | A | 17 | 28 | 0.6071 | 2.689 | 0.1011 |
| 12 | rs812826 | 56495306 | T | C | 18 | 28 | 0.6429 | 2.174 | 0.1404 |
| 12 | rs773121 | 56498241 | A | G | 17 | 27 | 0.6296 | 2.273 | 0.1317 |

**Table S2**. PLINK transmission disequilibrium tests (TDT) based on affection status for the individual chromosome 12 SNPs.

| trait affection; offset 0.000; model additive; test bi-allelic; minsize 10; min_freq 0.000; p 1.000; maxcmh 1000 | | | | | | | | | | | | | | | | | | | | |
| --- | --- | --- | --- | --- | --- | --- | --- | --- | --- | --- | --- | --- | --- | --- | --- | --- | --- | --- | --- | --- |
| Haplotypes and EM estimates of frequency | | | | | | | | | | | | | | | | | | | | |
|  | rs67594137 | | rs11171713 | | | rs11550558 | | rs7963590 | | rs12817471 | | rs2229046 | | rs773123 | | rs812826 | | rs773121 | |  |
| a1 | 1 | | 1 | | | 1 | | 1 | | 1 | | 1 | | 1 | | 1 | | 1 | | 0.823 |
| a2 | 2 | | 2 | | | 2 | | 2 | | 2 | | 2 | | 2 | | 2 | | 2 | | 0.093 |
| a3 | 1 | | 1 | | | 1 | | 1 | | 1 | | 1 | | 2 | | 2 | | 2 | | 0.048 |
| a4 | 1 | | 1 | | | 1 | | 1 | | 1 | | 1 | | 2 | | 1 | | 1 | | 0.012 |
| a5 | 1 | | 1 | | | 1 | | 1 | | 1 | | 1 | | 2 | | 2 | | 1 | | 0.004 |
| a6 | 2 | | 2 | | | 2 | | 2 | | 2 | | 2 | | 1 | | 1 | | 1 | | 0.004 |
| a7 | 2 | | 2 | | | 2 | | 1 | | 2 | | 2 | | 2 | | 2 | | 2 | | 0.004 |
| a8 | 1 | | 1 | | | 2 | | 1 | | 1 | | 1 | | 1 | | 1 | | 1 | | 0.004 |
| a9 | 2 | | 1 | | | 1 | | 1 | | 1 | | 1 | | 1 | | 1 | | 1 | | 0.004 |
| a10 | 1 | | 1 | | | 1 | | 2 | | 2 | | 2 | | 2 | | 2 | | 2 | | 0.004 |
| **Allele** | | **afreq** | | **fam#** | **S-E(S)** | | **Var(S)** | | **Z** | | **P** | |  | |  | |  | |  | |
| a1 | | 0.823 | | 33 | 5 | | 12.805 | | 1.397 | | 0.162329 | |  | |  | |  | |  | |
| a2 | | 0.093 | | 21 | -2 | | 7.233 | | -0.744 | | 0.457096 | |  | |  | |  | |  | |
| a3 | | 0.048 | | 11 | -3.5 | | 3.417 | | -1.894 | | 0.058291 | |  | |  | |  | |  | |

**Table S3**. Family-Based Association Tests (FBAT) HBAT test based on affection status for chromosome 12 SNPs.

| **SNP** | **PHRED** | **ConsDetail** | **GeneID** | **FeatureID** | **GeneName** |
| --- | --- | --- | --- | --- | --- |
| rs67594137 | 0.777 | non_coding_exon | ENSG00000237493 | ENST00000552016 | RP11-603J24.7 |
| rs67594137 | 0.777 | intron | ENSG00000111540 | ENST00000360299 | RAB5B |
| rs11171713 | 6.66 | non_coding_exon | ENSG00000237493 | ENST00000552016 | RP11-603J24.7 |
| rs11171713 | 6.66 | regulatory | NA | ENSR00001032243 | NA |
| rs11171713 | 6.66 | intron | ENSG00000111540 | ENST00000360299 | RAB5B |
| rs11550558 | 13.22 | upstream | ENSG00000139531 | ENST00000266971 | SUOX |
| rs11550558 | 13.22 | non_coding_exon | ENSG00000255990 | ENST00000541217 | AC034102.1 |
| rs11550558 | 13.22 | 3_prime_UTR | ENSG00000111540 | ENST00000360299 | RAB5B |
| rs7963590 | 8.792 | regulatory | NA | ENSR00001032245 | NA |
| rs7963590 | 8.792 | 5_prime_UTR | ENSG00000139531 | ENST00000394109 | SUOX |
| rs12817471 | 0.865 | regulatory | NA | ENSR00000268565 | NA |
| rs12817471 | 0.865 | intron | ENSG00000065361 | ENST00000267101 | ERBB3 |
| rs2229046 | 9.385 | synonymous | ENSG00000065361 | ENST00000267101 | ERBB3 |
| rs773123 | 24.6 | missense | ENSG00000065361 | ENST00000267101 | ERBB3 |
| rs773123 | 24.6 | downstream | ENSG00000257553 | ENST00000548595 | RP11-603J24.17 |
| rs773123 | 24.6 | upstream | ENSG00000170515 | ENST00000303305 | PA2G4 |
| rs773123 | 24.6 | upstream | ENSG00000257411 | ENST00000548861 | RP11-603J24.9 |
| rs812826 | 8.737 | downstream | ENSG00000257553 | ENST00000548595 | RP11-603J24.17 |
| rs812826 | 8.737 | upstream | ENSG00000170515 | ENST00000303305 | PA2G4 |
| rs812826 | 8.737 | intron | ENSG00000257411 | ENST00000548861 | RP11-603J24.9 |
| rs812826 | 8.737 | splice,intron | ENSG00000065361 | ENST00000267101 | ERBB3 |
| rs773121 | 13.2 | downstream | ENSG00000065361 | ENST00000267101 | ERBB3 |
| rs773121 | 13.2 | downstream | ENSG00000257553 | ENST00000548595 | RP11-603J24.17 |
| rs773121 | 13.2 | regulatory | NA | ENSR00000052477 | NA |
| rs773121 | 13.2 | 5_prime_UTR | ENSG00000170515 | ENST00000303305 | PA2G4 |
| rs773121 | 13.2 | intron | ENSG00000257411 | ENST00000548861 | RP11-603J24.9 |

**Table S4**. CADD PHRED scores and a subset of annotation details for chromosome 12 SNPs.

|  |  | **Minor Allele Frequency** | | |
| --- | --- | --- | --- | --- |
| **Gene** | **SNP** | **European** | **East Asian** | **African American** |
| *ERBB3* | rs7297175 | 0.5963 | 0.7735 | 0.5060 |
| *ERBB3* | rs12817471 | 0.0746 | 0.0000 | 0.0785 |
| *ERBB3* | rs2229046 | 0.0764 | 0.0090 | 0.0779 |
| *ERBB3* | rs773123 | 0.1101 | 0.0011 | 0.0836 |
| *ERBB3* | rs812826 | 0.1103 | 0.0011 | 0.1577 |
| *ERBB3* | rs773121 | 0.1055 | 0.0006 | 0.1597 |
| *RAB5B* | rs67594137 | 0.0791 | 0.0039 | 0.0850 |
| *RAB5B* | rs11171713 | 0.0773 | 0.0039 | 0.0788 |
| *RAB5B* | rs11550558 | 0.0780 | 0.0122 | 0.3545 |
| *SUOX* | rs7963590 | 0.7796 | 0.0000 | 0.0610 |

**Table S5. Minor Allele Frequencies for the 12q13.2** **SNPs.** Minor allele frequencies for non-Finnish Europeans, East Asians and African Americans were extracted from GnomAD (<https://gnomad.broadinstitute.org>). rs7297175 is independent of the other 9 SNPs and is not included in either haplotype block.
